# Predicting the impact of disruptions in lymphatic filariasis elimination programmes due to the outbreak of coronavirus disease (COVID-19) and possible mitigation strategies

**DOI:** 10.1101/2020.11.06.20227017

**Authors:** Joaquín M. Prada, Wilma A. Stolk, Emma L. Davis, Panayiota Touloupou, Swarnali Sharma, Johanna Muñoz, Rocio M. Caja Rivera, Lisa J. Reimer, Edwin Michael, Sake J de Vlas, T Déirdre Hollingsworth

**Affiliations:** School of Veterinary Medicine, Faculty of Health and Medical Sciences, University of Surrey, Guildford, UK; Department of Public Health, Erasmus MC, University Medical Center Rotterdam, Rotterdam, The Netherlands; Big Data Institute, Li Ka Shing Center for Health Information and Discovery, Headington, Oxford, UK; Department of Statistics, University of Warwick, Coventry, UK; Department of Biological Sciences, University of Notre Dame, South Bend, Indiana, USA; Department of Vector Biology, Liverpool School of Tropical Medicine, L35QA, UK

**Keywords:** Acceleration, COVID-19, Elimination, Lymphatic filariasis, Mitigation, Modelling

## Abstract

**Background:** In view of the current global COVID-19 pandemic, mass drug administration interventions for neglected tropical diseases, including lymphatic filariasis, have been halted. We used mathematical modelling to estimate the impact of delaying or cancelling treatment rounds and explore possible mitigation strategies.

**Methods:** We used three established lymphatic filariasis transmission models to simulate infection trends in settings with annual treatment rounds and programme delays in 2020 of 6, 12, 18 or 24 months. We then evaluated the impact of various mitigation strategies upon resuming activities.

**Results:** The delay in achieving the elimination goals is on average similar to the number of years the treatment rounds are missed. Enhanced interventions implemented for as little as one year can allow catch-up on the progress lost, and if maintained throughout the programme can lead to acceleration of up to 3 years.

**Conclusions:** In general, a short delay in the programme does not cause major delay in achieving the goals. Impact is strongest in high endemicity areas. Mitigation strategies such as biannual treatment or increased coverage are key to minimizing the impact of the disruption once the programme resumes; and lead to potential acceleration, should these enhanced strategies be maintained.

## Introduction

Lymphatic filariasis (LF), a disease caused by parasitic filarial worms, was identified as potentially eradicable in 1993^1^. It has been targeted for elimination as a public health problem by the World Health Organization (WHO), with initial goals set for 2020, which are now being revised for 2020^2^. The main strategy to achieve elimination as a public health problem is through mass drug administration (MDA). The most common drug combinations used by the LF control and elimination programmes is either a combination of diethylcarbamazine citrate and albendazole (DA), or ivermectin and albendazole (IA) in areas where onchocerciasis is endemic. Geographically, this means that IA is used in most of Africa, while DA is used in other parts of the world, such as the Indian subcontinent. Currently, MDA programmes are ongoing in 46 countries^3^.

Mass drug administration campaigns are generally carried out annually, for a minimum of five years (5 rounds), with the aim of achieving at least 65% coverage. Afterwards, a series of transmission assessment surveys must reveal low likelihood of current transmission to achieve elimination as a public health problem. This is generally associated to reaching a threshold of 1% microfilaria (mf) prevalence, or 2% antigenemia prevalence, in areas where the dominant vector for transmission is *Anopheles* or *Culex* (0.5% mf threshold used in areas with *Aedes*-transmitted LF)^4^.

The current global COVID-19 pandemic has had a huge impact world-wide, with over 10 million confirmed cases in the first half of 2020 and over half a million deaths. This impact is compounded by its indirect effects on other diseases and health programmes. In view of the current global COVID-19 pandemic, and the need to practice physical distancing, WHO issued the recommendation on 1 April 2020 to put all community-based surveys, active case-finding activities and mass treatment campaigns for neglected tropical diseases on hold until further notice^5^. For the LF control and elimination programmes which had an MDA round planned, it has meant a stop of all activities. It thus becomes important to assess the impact of delays in the MDA delivery on the 2030 goals, and to consider strategies to strengthen programmes after the lockdown to mitigate the negative impact of disruption.

Strategies have been considered for accelerating progress towards the 2020 goals (now 2030) which could be used as mitigation strategies, such as biannual rounds of treatment or increased coverage^6–8^. Recent clinical trials and mathematical modelling studies have shown that the combination of all three drugs, commonly known as the triple drug (IDA), can substantially improve progress towards the goal ^7,9–13^. The use of IDA has been recommended by WHO to accelerate global elimination efforts^14^. However, only countries without onchocerciasis can deploy IDA in their programmes, due to risks of adverse events.

A suite of three different mathematical models have been recently used to compare strategies to accelerate global elimination of LF^7^, as well as to assess the likelihood of resurgence after reaching the 1% mf prevalence threshold^8^. Those same models are used here to: 1) estimate the impact of MDA delays in the expected progress towards the 1% mf threshold, 2) establish mitigation strategies that are sufficient to catch-up the missed/delayed rounds, and 3) assess the acceleration provided from mitigation strategies that can be maintained longer term. Mitigation and acceleration strategies considered here include enhancing coverage from 65% to 80%, deploying the treatment twice per year (biannual treatment), and, for areas using DA, the benefit of switching to the triple drug.

## Materials and Methods

### Employed mathematical models

We used three well-described published mathematical models for LF transmission to enhance our understanding of the disruption caused by COVID-19 to ongoing control and elimination efforts. We included the following models, developed and applied by members of the Neglected Tropical Disease Modeling Consortium: EPIFIL^15,16^, a deterministic population-based model; and LYMFASIM^17,18^ and TRANSFIL^19,20^, both stochastic individual based models. All models capture the basic processes relevant to the transmission dynamics of LF, including parasite life cycle, vector density and biting rate, and human exposure to the vectors. The formulation and parametrization of these models is detailed in the references above, see supplementary material for an updated implementation of the three models.

### Scenarios considered

Countries may be at different stages in their control programme. In this analysis, we assumed that two annual MDA rounds achieving 65% coverage were completed before the interruption caused by COVID-19 (disruptions at other points in the programme yield broadly similar results and are discussed in the supplementary material, page 2). Two settings were considered, one relevant for most of Africa, with *Anopheles*-driven transmission and annual treatment with IA; and a second representing India-like populations, with *Culex*-driven transmission and treatment with DA. The assumptions of drug effectiveness, in terms of macro and microfilaricidal effect and sterilization of adult worms, was taken from Stolk, Prada et al.^7^, see also supplementary material (page 4). Vector control is commonly recommended to enhance MDA, but is generally outside the control of the LF programmes. To be conservative, we considered that there was no bednet coverage, while we acknowledge that bednet coverage is present in many African areas^21^. Additional simulations considering bednets and indoor residual spraying (IRS) are shown in the supplementary materials (page 3).

To assess the impact of delayed MDA, we explored four scenarios where the MDA rounds are postponed either 6, 12, 18 or 24 months, which causes a gap of one and a half, two, two and a half, and three years respectively, Figure 1. A 6 month interruption represents one postponed round and 12 or 24 month interruptions would represent one or two cancelled rounds respectively, whereas an 18 month interruption represents one delayed and one postponed round.

**Figure 1.**
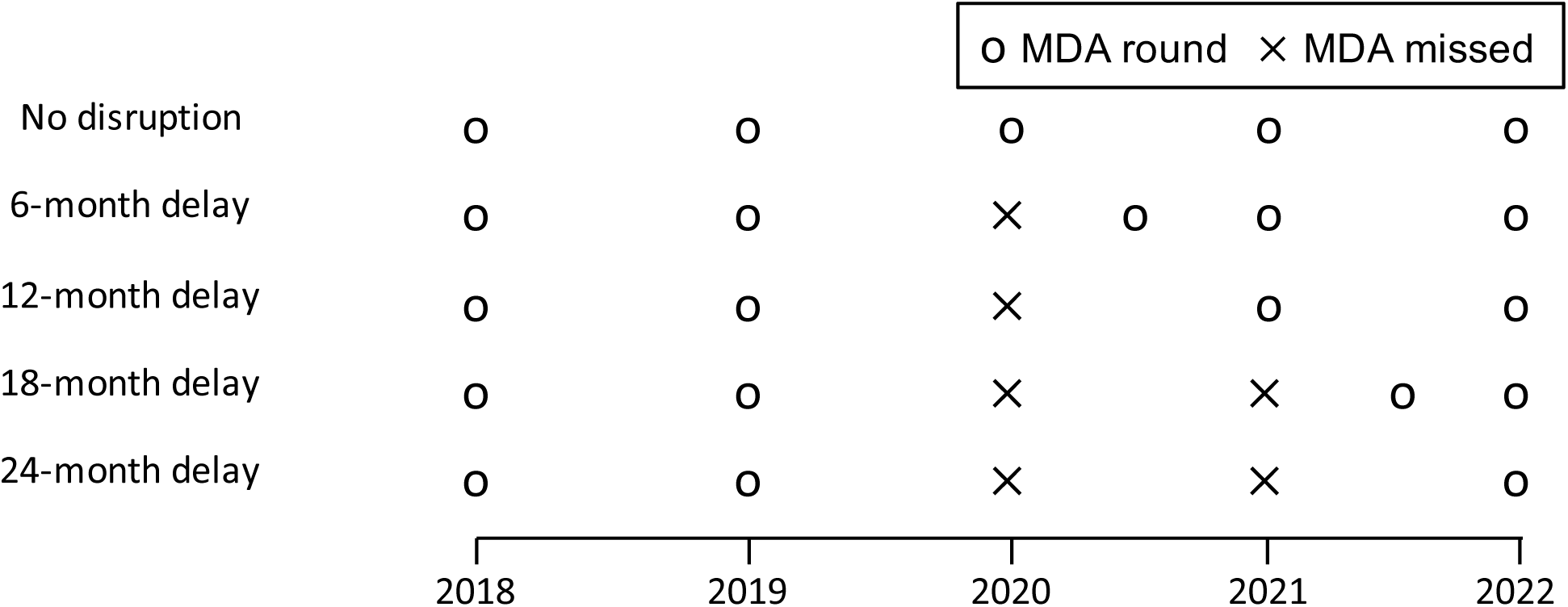
Timing of the MDA rounds assuming the programme starts in 2018, and the COVID-19 disruption occurs in 2020. Four delay scenarios (6, 12, 18 and 24 months) and a no disruption scenario were considered. A treatment round is merely postponed if it is delayed for 6 months, but is cancelled if delayed for 12 months (resulting in 1 cancelled round in the 12- and 18-months delay scenarios, and two cancelled rounds in the 24-months delay scenario).

To manage and minimize the impact of the delay in delivering the next round of MDA, national programmes could implement alternative enhanced strategies after resuming activities. Here, we explored the impact of various MDA-based mitigation strategies, namely i) enhanced achieved coverage (up from 65% to 80%); ii) switching to biannual rounds, treating every six months; and iii) in areas where appropriate (here our India-like setting), the deployment of the triple drug. In all simulated scenarios, MDA continues until the 1% mf threshold is achieved, up to 2045 (end of simulation window).

### Analysis of simulation results

We generated a large number of simulations (simulation bank) and extracted uniformly 10,000 runs per scenario, for each model, across a wide range of baseline prevalence (assumed to be in 2018), from 1% to 40%. All simulations across the three models are considered as an ensemble, with equal weighting. We projected forward from the baseline under the different treatment regimes in each scenario and extracted the simulated mf prevalence in the population, each year. We summarized some results following a general classification of low, medium and high prevalence areas, these were mapped to the non-contiguous microfilaria prevalence of 5-10%, 15-20% and 25-30% respectively. To select the simulations for each prevalence bracket, we extracted 1,000 runs from each model uniformly from the simulation bank.

We used a counterfactual no disruption scenario (i.e. the initial five rounds take place as expected, Figure 1), to assess for how long the enhanced strategies need to be implemented in order to catch up with the expected progress of the programme. We calculated the year in which the prevalence in the different interventions is equal or below the counterfactual scenario, as that would indicate that the enhanced strategy managed to mitigate the disruption caused by the gap in treatment, see Figure 2 for an illustrative example. After the initial 5 year programme, we assume that the treatment strategy used after resuming activities is continued until the 1% mf threshold is met. We can then use these same runs to quantify the gains should these alternative strategies be maintained beyond catching up, see Figure 2. To summarize results over the prevalence range, we calculated the moving average with a window size of 4,000 and the volatility as the unweighted standard deviation in the same window size.

**Figure 2.**
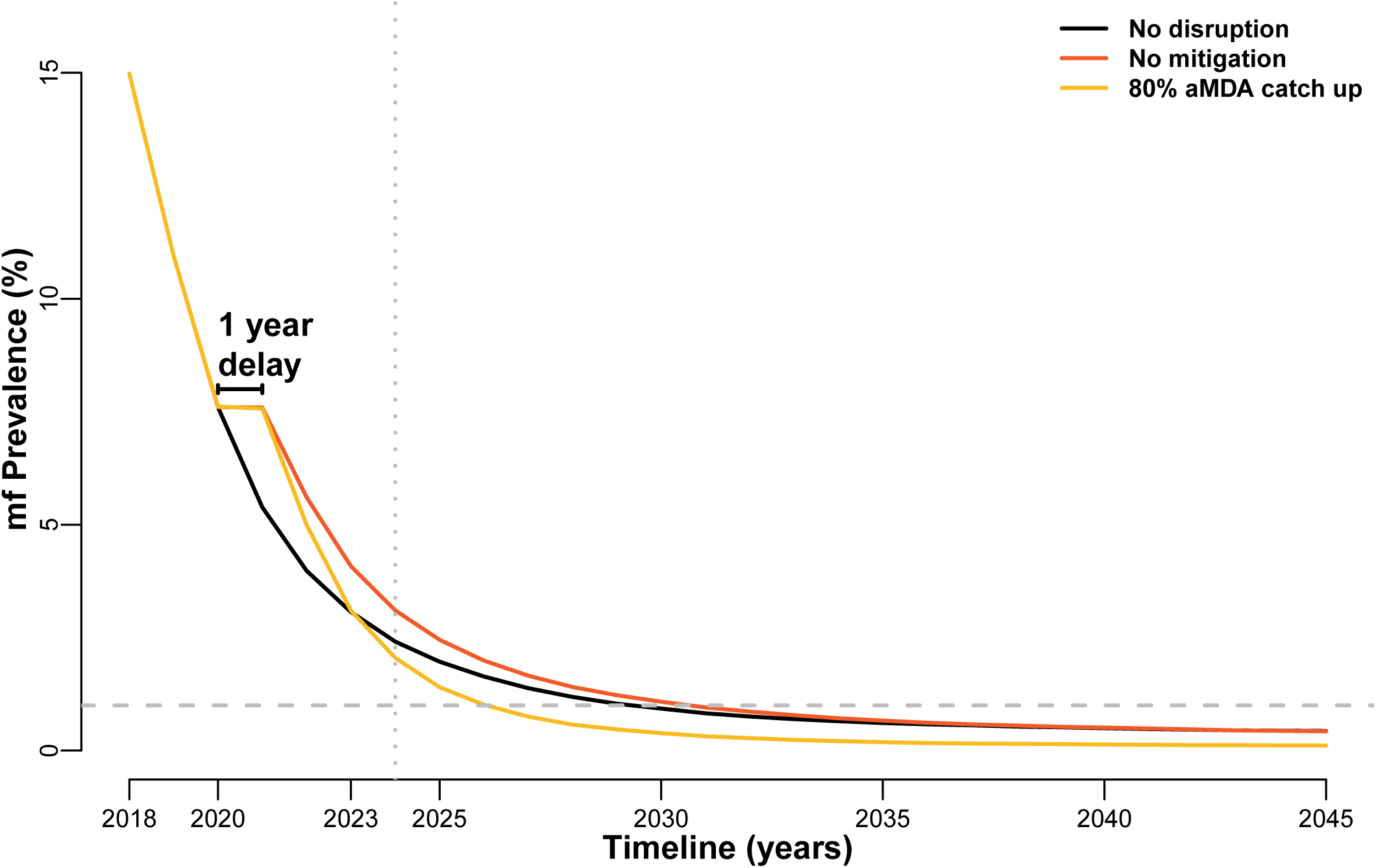
Example of yearly mf prevalence trends over time. Black line shows a non disruption scenario (counterfactual), with a baseline prevalence of 15%. Red line shows a no mitigation scenario, where a one year delay takes place in 2020. The yellow line shows a scenario where 80% aMDA is implemented from 2021 onward as a catch-up (and acceleration) strategy. Vertical dotted line indicates the catch-up point (the first year where the yellow line is below the black line). Horizontal dashed line marks the 1% mf prevalence threshold, which is reached earlier in the acceleration (yellow) scenario. In this example it takes 3 rounds of aMDA with 80% coverage to catch-up. All three solid lines are an average of 1,000 simulations with TRANSFIL, with a baseline prevalence in 2018 between 14% and 16%.

## Results

The expected delay in reaching the 1% mf threshold is broadly similar across the baseline prevalence (here considered in 2018), Figure 3 - top. Across the full baseline prevalence range explored, the delay in reaching the 1% goal (and thus the increase in the length of the programme that needs to be considered) is estimated to be on average slightly less than the delay of the MDA round. For example, a one year delay in distributing the MDA round causes on average almost one year delay in the *Culex*, DA setting and a bit less than a one year delay in the *Anopheles*, IA setting, Figure 3 - top. Areas with prevalence below 5% are close to the goal, and thus the disruption is minimal. The proportion missing the 2030 goal is estimated to increase faster in *Anopheles* settings than in *Culex* settings as baseline prevalence increases, Figure 3 - bottom.

**Figure 3.**
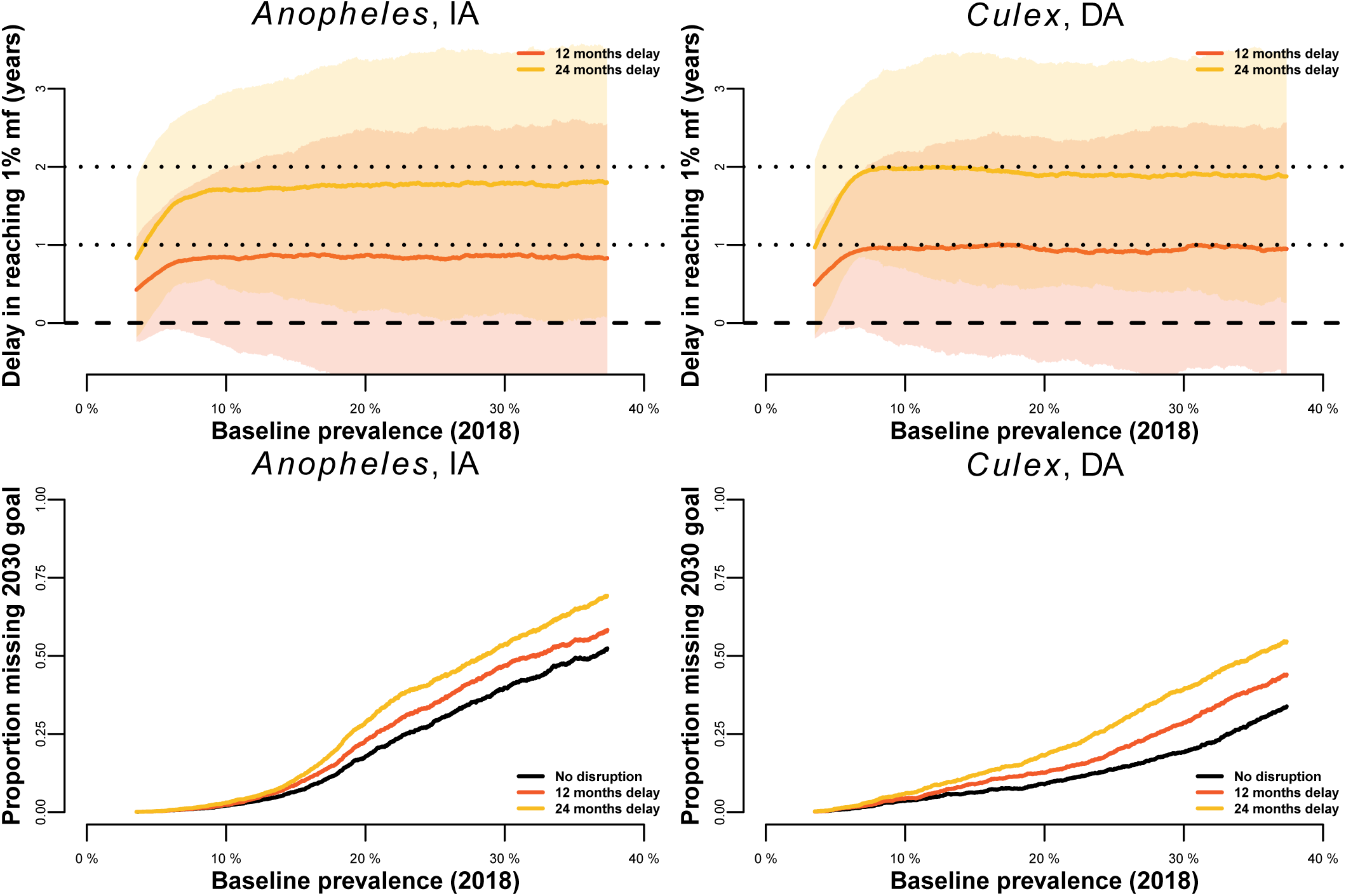
Moving average (window size of 4,000) of the delay in reaching 1% mf prevalence (in years - top), and proportion missing the 2030 goals (bottom), for a wide range of baseline prevalence at the start of the current MDA programme in an *Anopheles*-transmitted setting treating with IA (left) and a *Culex*-transmitted setting treating with DA (right). The red and yellow lines illustrate scenarios with a 12 or 24 months programme delay respectively, after which annual MDA with 65% coverage continues. Shaded colour areas illustrate the standard deviation (volatility).

A summary of the results across the four delays we considered, for the two settings, is shown in Table 1. The focus here are the low, medium and high prevalence areas, as defined above (5-10%, 15-20% and 25-30% mf prevalence respectively). Low prevalence areas, irrespective of the delay considered, are on average likely to reach 1% mf before the 2030 endpoint, however delays in medium and high prevalence areas may require programmes to continue beyond 2030. Across the two settings and the prevalence brackets considered, a 6 month delay in the deployment of the MDA will lead on average to the programme not needing to be extended beyond the originally planned timelines, Table 1.

**Table 1.** Estimated timeline to achieving 1% mf prevalence goal by baseline endemicity, mean in years (95%CI), and estimated extension of the programmes required (i.e. delay to reaching 1% mf) for different delays in the deployment of the next MDA. Timeframes over 12 years from the baseline are beyond 2030, and thus are estimated to miss the goal.

|  | Africa-like population<br>*Anopheles* transmission, IA treatment |  |  | India-like population<br>*Culex* transmission, DA treatment |  |  |
| --- | --- | --- | --- | --- | --- | --- |
| **Baseline prevalence (2018)** | Low Prevalence (5-10% mf) | Medium Prevalence (15-20% mf) | High Prevalence (25-30% mf) | Low Prevalence (5-10% mf) | Medium Prevalence (15-20% mf) | High Prevalence (25-30% mf) |
| **Time to goal from 2018 if no interruption (counterfactual )** | 5.89 years (3-9) | 9.51 years (7-18) | 11.8 years (8-21) | 5.57 years (4-12) | 8.54 years (6-17) | 10.13 years (7-19) |
| **6 month delay (1.5 year gap)** | no extension (0-2) | no extension (0-4) | no extension (0-4) | no extension (0-2) | no extension (0-3) | no extension (0-3) |
| **12 month delay (2 year gap)** | + 0.79 years (0-2) | + 0.81 years (0-4) | + 0.85 years (0-5) | + 0.96 years (0-3) | + 0.95 years (0-4) | + 0.92 years (0-4) |
| **18 month delay (2.5 year gap)** | + 0.7 years (0-2) | + 0.92 years (0-4) | + 1 years (0-5) | + 0.77 years (0-3) | + 0.97 years (0-4) | + 0.94 years (0-4) |
| **24 month delay (3 year gap)** | + 1.63 years (0-3) | + 1.72 years (0-5) | + 1.82 years (0-5) | + 1.92 years (0-4) | + 1.87 years (0-5) | + 1.85 years (0-5) |

Alternative enhanced strategies, such as increasing efforts to achieve a higher coverage or increasing the treatment frequency can help catch-up to the time lost due to the COVID-19 disruption. Biannual rounds of MDA or switching to IDA, where possible, are on average faster methods to catch up than achieving a high coverage of 80%, Figure 4. Longer gaps in the programme would require more rounds of the enhanced campaigns to catch up, see supplementary materials (page 5).

**Figure 4.**
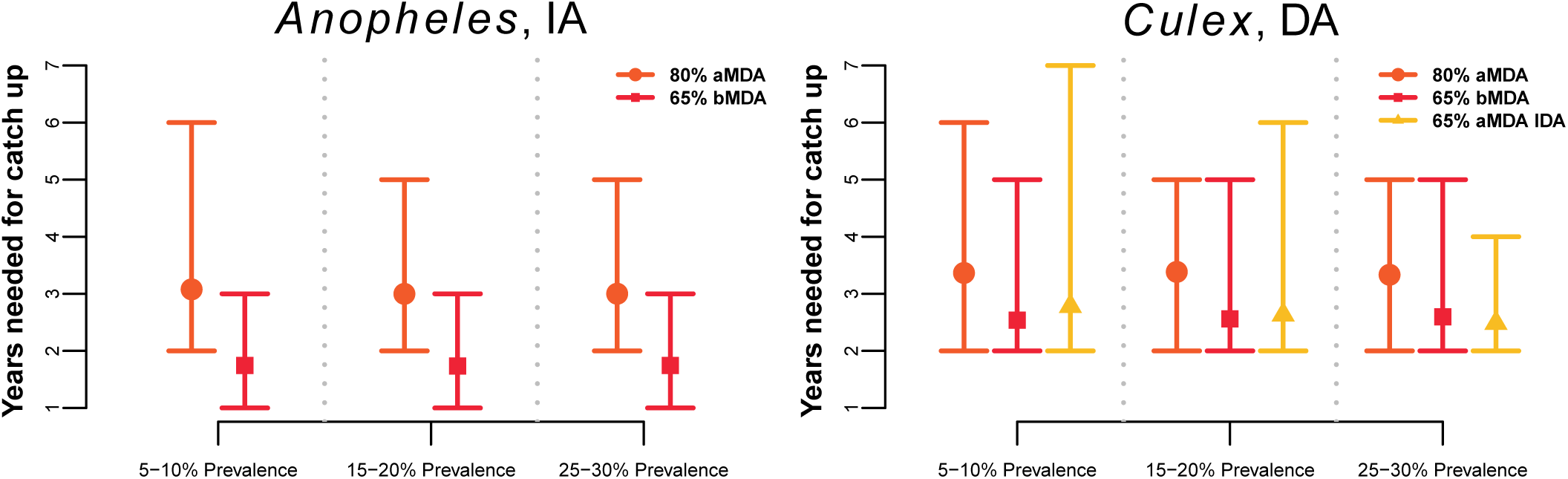
Estimated average number of years of the enhanced interventions needed, after a one year delay in aMDA and resuming activities, to catch-up to the counterfactual no disruption scenario, for the two settings considered.

Maintaining the strategies implemented to catch-up and mitigate the disruption caused by COVID-19 will lead to acceleration of the programmes, irrespective of the strategy used, Figure 5. A long disruption causing a two years delay (a three year gap between treatment rounds), can be caught up by these alternative strategies. All three strategies considered here (80% coverage achieved, biannual rounds or switching to IDA where possible), lead to qualitatively similar gains, Figure 5, which could be as big as reaching the goal three years earlier. The results shown here are for an India-like setting, with *Culex* as the dominant vector and the deployment of DA as the drug combination, results for *Anopheles*-transmitted setting, treated with IA, are broadly similar, see supplementary material (page 6).

**Figure 5.**
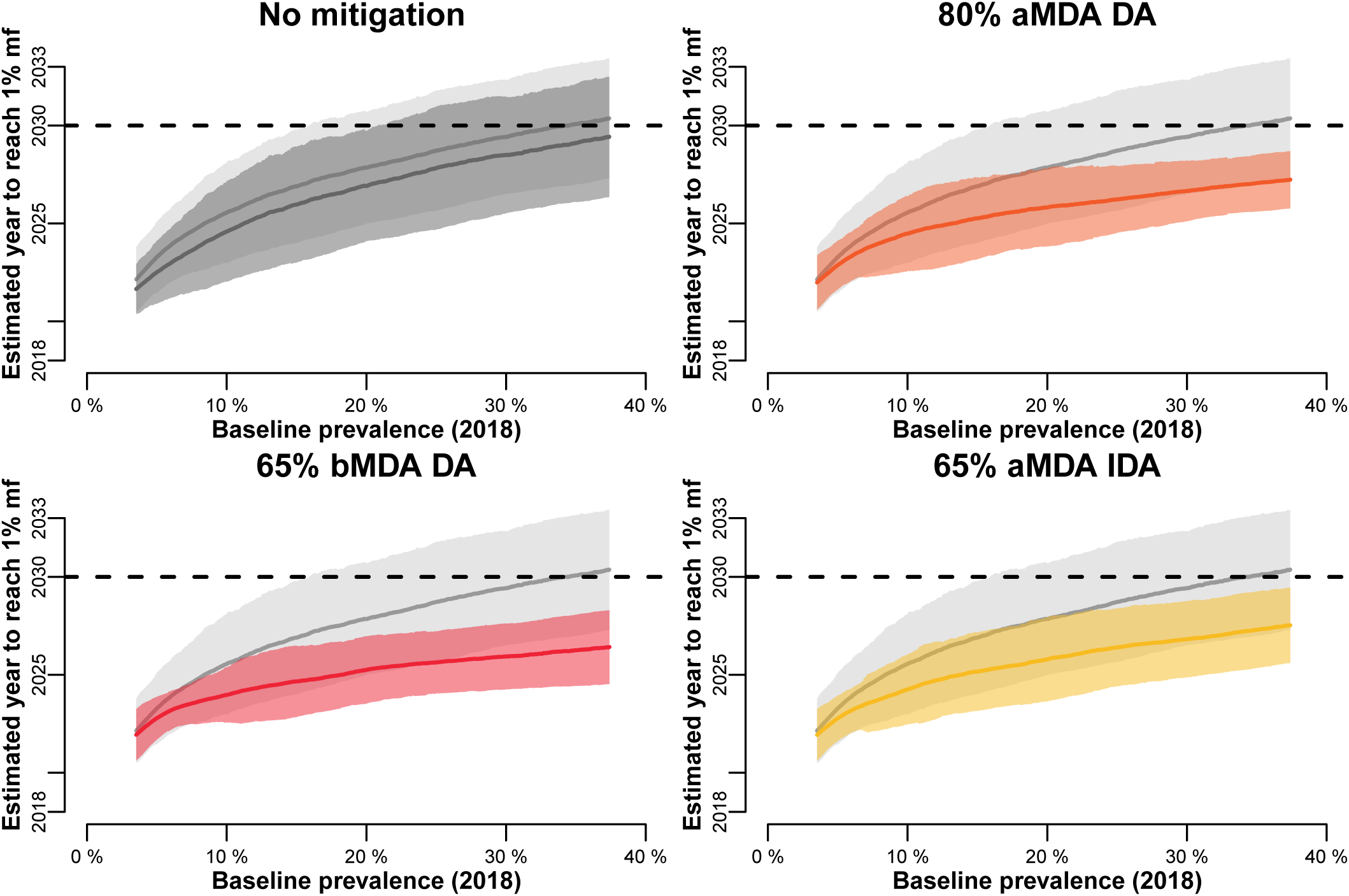
Estimated year of achieving the 1% mf threshold, relative to the prevalence at baseline (assumed here in 2018), for setting with *Culex* transmission treated with DA. Top-left plot compares the counterfactual to the no mitigation scenario (after a 12 month delay). The rest of the plots show the different acceleration strategies (in colour) against the no mitigation situation (in grey). Horizontal dashed line indicates the year 2030, areas that go above 2030 are unlikely to reach the goal. Solid line shows the moving average (window size of 4000), shaded area is the standard deviation (volatility).

## Discussion

We have estimated the delays in LF elimination efforts due to the disruption caused by COVID-19, in two broadly defined settings, using three well-established mathematical models of lymphatic filariasis transmission^7,8^. Moreover, we explored how alternative enhanced strategies could be used to catch-up and recover from the disruption when programmes resume, and potentially accelerate progress towards the 2030 goals, if maintained longer term.

Overall, our simulations suggest that each round missed generally extends the programme by the same amount, Figure 3, provided programmes can resume activities achieving similar levels of coverage as before the disruption. As an example, a programme that misses one round this year, and resumes their MDA campaigns next year, is likely to be one year behind, although in some cases it could lead to longer delays, Figure 3 / Table 1. Moreover, baseline prevalence has a very limited effect on the delays, as seen in Figure 3, with the exception of very low prevalence areas (<5% mf). However, the estimated time to reach 1% mf prevalence has a larger uncertainty in high prevalence areas, Table 1. These results are consistent across the two settings considered (*Anopheles* dominated transmission with IA-based MDA and *Culex* dominated transmission with DA-based MDA).

Medium (15-20% mf), and high (25-30% mf) prevalence areas are more at risk of missing the 2030 target, should the delays not be mitigated. In low prevalence areas (as defined above, 5-10% mf in 2018), a two-year delay (three-year gap between treatment rounds), could lead in some extreme cases to missing the 2030 goal, Table 1. However, we expect that on average those areas, or those with even lower baseline prevalence, will reach the 2030 goals ahead of time, in spite of the COVID-19 disruption. In page 2 of the supplementary materials we show that results for programmes that started earlier/later their MDA are qualitatively similar.

Encouragingly, interventions that are commonly considered to accelerate progress towards the goals, such as increased coverage or frequency of the interventions^7^, can also be used to mitigate the impact of the missed rounds. A 6-month delay in the MDA programme, effectively deploying two MDA rounds 6 months apart, see Figure 1, is similar to resuming activities with a biannual strategy, where one year is sufficient to catch up to the expected progress of the programme in the absence of any delays, Figure 4. These results highlight the importance of implementing the missed round as soon as possible when it is safe to do so, which would also prevent the expiry of medication already stocked. Should these enhanced strategies be maintained, once the programmes resume activities, until the 1% mf threshold is reached, as much as three years could be gained, Figure 5. Our models predict that the extended use of these enhanced strategies will ensure that even high prevalence areas will be able to reach the 2030 targets.

Our analysis has a number of assumptions and simplifications that need to be considered. While we allow parameter uncertainty in the inputs (see supplementary material), and for LYMFASIM and TRANSFIL, stochastic effects, we do not account for sampling of the human population, and thus the mf is the true value in the population over 5 years (minimum age to be included in a survey). Similarly, the deployment of the drug and its effect takes place within one time-step of the model (two weeks/one month depending on the model). This, combined with the assumption that compliance (the proportion of individuals treated in two consecutive rounds) is the same between biannual rounds as between annual rounds, means that our results for biannual treatment might be slightly optimistic. We have conducted a sensitivity analysis for other simplifications, such as the effect of vector control and the disruption occurring at a different point in the programme, see supplementary materials (pages 2-3).

For our analysis, we have assumed that the programmes can resume MDA activities with a similar coverage as before the disruption. In previous occasions where MDA has been interrupted, programmes have reported good coverage the following years after resuming activities. In Haiti, following the 2010 earthquake, there was a 92% reported coverage of MDA for LF in 2011, which was calculated from the doses administered and estimated population sizes; a household survey in Port-au-Prince the same year reported a coverage of 71%^22^. Similarly, the Sierra Leone and Liberia LF programmes missed MDA in 2014 due to the West Africa Ebola epidemic^23^, but both managed to resume in 2015 and report >70% coverage^24,25^. However, whilst Guinea reported a 16% coverage achieved during the outbreak, this only increased to 21% in 2015; while by 2016 coverage was reported up to 73%^26^. Unfortunately, there is little information available on how these interruptions affected program outcomes, as the majority are still undertaking MDA, but by 2019, 9 of 14 districts in Sierra Leone had stopped MDA^27^.

One important aspect to consider, which we currently do not capture in our models, is that a delay in meeting the 1% mf elimination as a public health threshold could cause higher morbidity levels while transmission is ongoing. This may lead to new incident cases of morbidity, especially in areas that have not started MDA yet or with a relatively recent start. Even when implementing catch up strategies, the time spent with a higher prevalence than originally planned will undoubtedly lead to higher morbidity in the affected communities. Moreover, morbidity management measures to relieve the suffering of those affected by hydrocele or lymphoedema may also be disrupted. Hydrocele operations will likely be delayed. Management of lymphoedema might perhaps continue, as it is commonly managed at home, although disruptions might make this more difficult. Therefore, it is not only important to mitigate the disruption, but also to do it quickly.

In summary, progress towards the LF 2030 goals is not going to be greatly affected by the COVID-19 disruption, if the interruption remains restricted to 6-24 months, especially if mitigation strategies are put in place. These enhanced strategies that will allow catching up are not particularly novel, and have been discussed and considered recently for accelerating progress towards the 2030 goals. An opportunity could be present for programmes that resume MDA using one of these mitigation strategies, particularly those that recently started, as it will lead to a faster reduction in microfilaria prevalence, and lower morbidity, should the strategies be maintained until the 1% mf threshold is met.

## Data Availability

No unpublished data was collected or used for this study. All additional information available in the supplementary materials

## Authors disclaimer

The funders had no role in study design, data collection and analysis, decision to publish, or preparation of the manuscript.

## Authors contributions

JMP, WAS, ELD, PT and TDH conceived the study. JMP designed the study, carried out the analysis, and drafted the manuscript. Model simulations were performed by SS and RMCR (EPIFIL), WAS and JM (LYMFASIM), and PT and ELD (TRANSFIL). All authors contributed to the interpretation of results and critically reviewed the manuscript.

## Funding

This work was supported by the Bill and Melinda Gates Foundation through the NTD Modelling Consortium (grant number OPP1184344).

## Competing interests

None declared

## Ethical Approval

Not required

## Notes

### Competing Interest Statement

The authors have declared no competing interest.

### Author Declarations

No unpublished data was collected or used for this study. Thus IRB approval was not needed.

