## Supplementary Materials for "Predicting the impact of disruptions in lymphatic filariasis elimination programmes due to the outbreak of coronavirus disease (COVID-19) and possible mitigation strategies"

### Supplementary Material

Prada *et al.* 2020

#### Index

| | |
|---|---|
| Sensitivity to timing of disruption | 2 |
| Sensitivity to vector control use | 3 |
| Treatment efficacy assumptions | 4 |
| Catch-up rounds needed with a 24 month delay | 5 |
| Acceleration to 2030 goals in *Anopheles*-transmitted setting | 6 |
| EPIFIL model description and methods | 7 |
| LYMFASIM model description and methods | 15 |
| TRANSFIL model description and methods | 29 |

#### Sensitivity to timing of disruption

We evaluated whether the disruption caused by COVID-19 would have a different impact in delaying reaching the 1% microfilaria threshold depending on how many previous rounds of treatment have taken place since the most recent baseline survey. We used the model TRANSFIL to simulate populations in a setting with Culex transmission treated with DA, and in a setting with Anopheles transmission treated with IA, and considered three possible timings for the disruption to happen:

1. As in the main text, the disruption happens mid-programme. We assume this translates to programmes that started in 2018 (most recent baseline survey), and in 2020 were expected to carry out the third treatment round.

2. The disruption happens at the end of the programme. We assume this translated to programmes that started in 2016 (most recent baseline survey), and in 2020 were expected to carry out the fifth (and last) treatment round.

3. The disruption happens at the start of the programme. We assume this translated to programmes that were planned to start in 2020 (most recent baseline survey), and thus were expected to carry out their first treatment round.

In all three timings above, the round due in 2020 is cancelled and the programmes resume one year later. Following the methodology in the main text, we extracted 10,000 simulations from the TRANSFIL model, uniformly, in the prevalence range between 1% and 40%. To summarize results over the prevalence range, we calculated the moving average with a window size of 2,000 and the volatility as the unweighted standard deviation in the same window size.

The results shown in Figure S1 suggest that the timing of the disruption does not affect the average delay of the programme. Note that for disruptions at the start of the programme, cancelling the first round, essentially translates to a one year delay. The uncertainty is however much wider in that scenario, as treatment rounds reduce the variation in mf prevalence across the simulations.

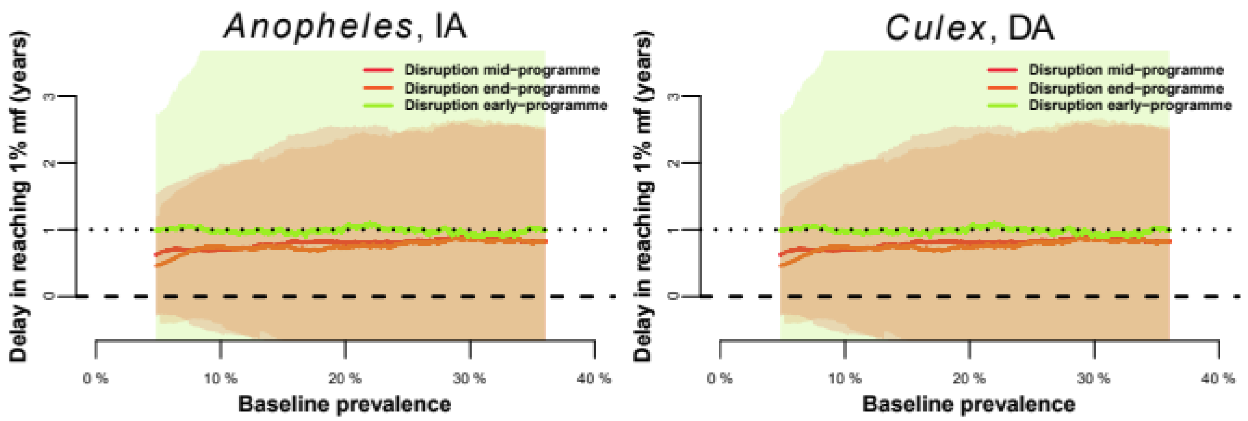

Figure S1. Moving average of the delay in reaching 1% mf prevalence (in years) for a wide range of baseline prevalence at the start of the current MDA programme. The red, orange and lime lines illustrate respectively a disruption mid-programme, at the end of the programme and at the beginning of the programme. Shaded colour areas illustrate the standard deviation (volatility).

#### Sensitivity to vector control use

We conducted additional simulations to evaluate the impact of vector control in the context of the disruptions explored in the main manuscript. We used the model EPIFIL to simulate populations in a setting with Culex transmission treated with DA and assumed that a bednet and an indoor residual spraying (IRS) coverage of 30% was achieved, and was maintained without disruption (even when MDAs are delayed). The implementation of vector control in the model is detailed below in the EPIFIL model description section. We explored two delays in MDA due to the disruption caused by COVID-19: a 12 month delay and a 24 month delay in the delivery of treatment.

Following the methodology in the main text, we extracted 10,000 simulations from the EPIFIL model, uniformly, in the prevalence range between 1% and 40%. To summarize results over the prevalence range, we calculated the moving average with a window size of 2,000 and the volatility as the unweighted standard deviation in the same window size.

The results shown in Figure S2 - left show the EPIFIL results similar to Figure 3 in the main text. Adding low level vector control can potentially reduce the average delays by around 15% and 32% for the 12 and 24 month delay scenarios respectively, Figure S2 – right. It is important to note the baseline in the counterfactual itself (i.e. no disruption scenario) will already change due to the inclusion of vector control, with a decreased prevalence, thus reducing delays by half a year on average (not shown).

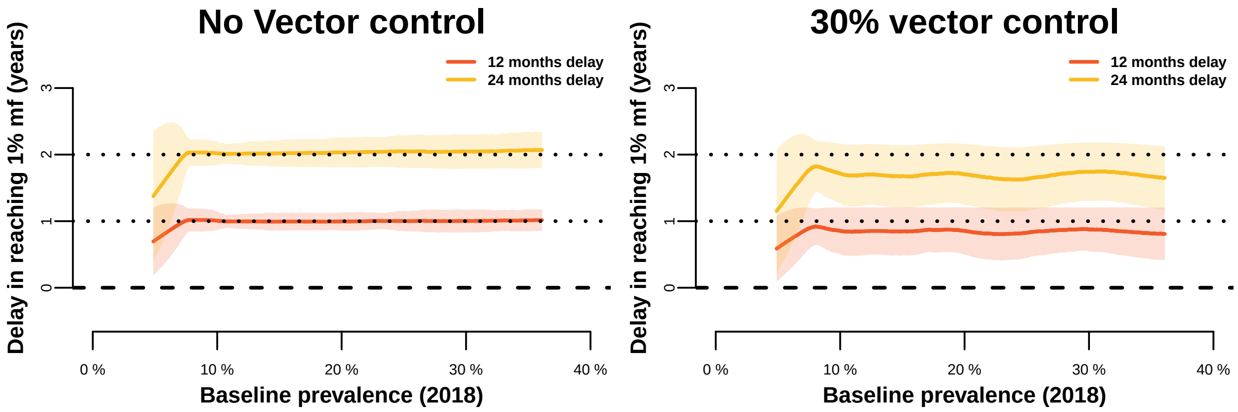

Figure S2. Moving average of the delay in reaching 1% mf prevalence (in years) for a wide range of baseline prevalence at the start of the current MDA programme in an a Culex-transmitted setting treating with DA, with vector control (right) and without (right). The red and yellow lines illustrate scenarios with a 12 or 24 months programme delay respectively; shaded colour areas illustrate the standard deviation (volatility).

#### Treatment efficacy assumptions

All 3 models used the same assumptions on treatment efficacy, with treatment regimens differing in the proportion of adult worms and mf killed, and a temporary or permanent reduction in mf productivity by surviving adult worms, following Stolk, Prada et al. (2018) as indicated in Table S1.

Table S1. Treatment efficacy assumptions adopted in the models

| Treatment Regimen | Proportion of Adult Worms Killed, % | Duration of Sterilization (months) | Proportion of Microfilariae Killed, % |
|---|---|---|---|
| DEC + ALB | 55% | 6 | 95 |
| IVER + DEC + ALB (optimistic)* | 55% | Permanent | 100 |
| IVER + ALB | 35% | 9 | 99 |

Abbreviations: ALB, albendazole; DEC, diethylcarbamazine; IVER, ivermectin.

*Triple-drug regimen, optimistic treatment efficacy assumptions were adopted.

Stolk, W.A., Prada, J.M., Smith, M.E., Kontoroupis, P., De Vos, A.S., Touloupou, P., Irvine, M.A., Brown, P., Subramanian, S., Kloek, M. and Michael, E., 2018. Are alternative strategies required to accelerate the global elimination of lymphatic filariasis? Insights from mathematical models. Clinical infectious diseases, 66(Supplement_4), pp.S260-S266.

### Catch-up rounds needed with a 24 month delay

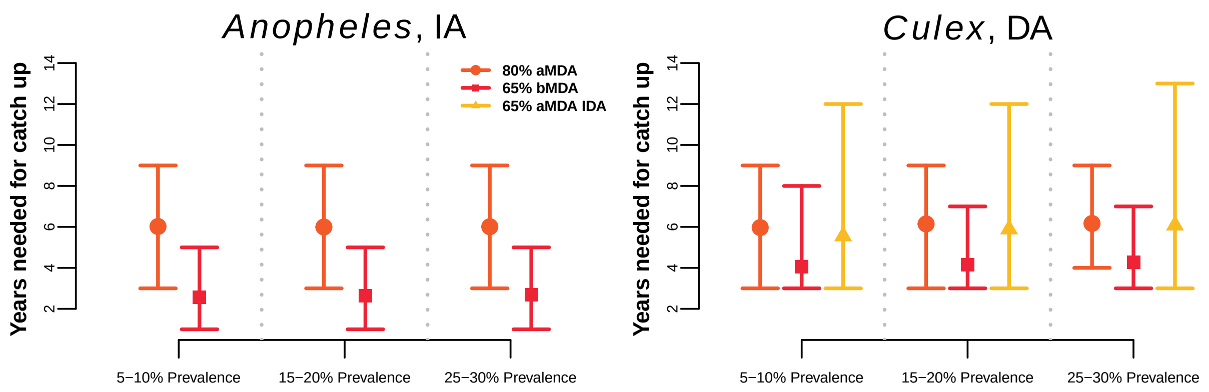

Figure S3. Estimated average number of years of the enhanced interventions needed, after a two year (24 month) delay in aMDA and resuming activities, to catch-up to the counterfactual no disruption scenario, for the two settings considered.

#### Acceleration to 2030 goals in *Anopheles*-transmitted setting

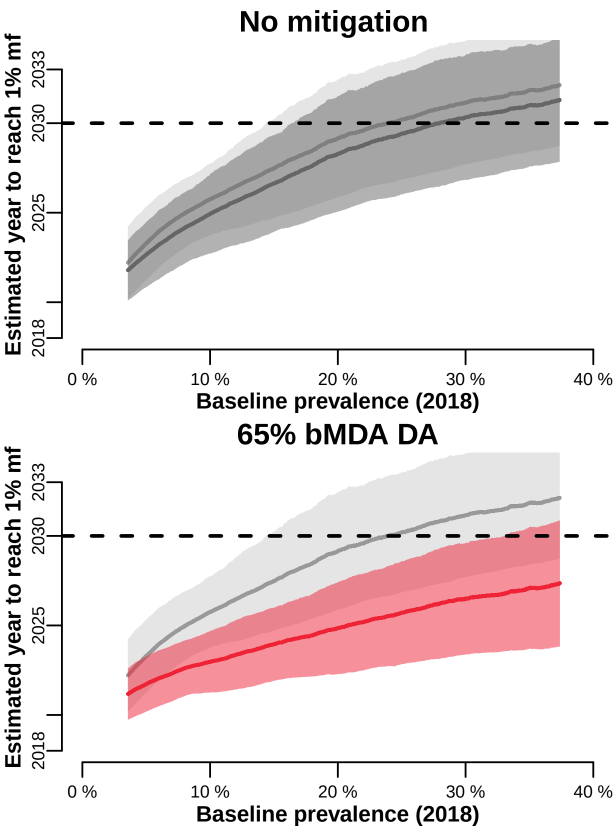

Figure S4. Estimated year of achieving the 1% mf threshold, relative to the prevalence at baseline (assumed here in 2018), for setting with *Anopheles* transmission treated with IA. Top-left plot compares the counterfactual to the no mitigation scenario (after a 12 month delay). The rest of the plots show the different acceleration strategies (in colour) against the no mitigation situation (in grey). Horizontal dashed line indicates the year 2030, areas that go above 2030 are unlikely to reach the goal. Solid line shows the moving average (window size of 4000), shaded area is the standard deviation (volatility).

#### EPIFIL model description and methods

##### The mathematical model of LF transmission dynamics

We employed a genus specific mosquito-vectored transmission model of LF to carry out the modelling work in this study [1-7]. Briefly, the state variables of this hybrid coupled partial differential and differential equation model vary over age (*a*) and/or time (*t*), representing changes in the pre-patent worm burden per human host $(P(a,t))$ adult worm burden per human host $(W(a,t))$ the microfilariae (mf) level in the human host modified to reflect infection detection in a 1 mL blood sample $(M(a,t))$ the average number of infective L3 larval stages per mosquito (*L*), and a measure of immunity $(I(a,t))$ developed by human hosts against L3 larvae. The state equations comprising this model are:

$$\frac{\partial P(a,t)}{\partial t}+\frac{\partial P(a,t)}{\partial a}=\lambda\frac{V}{H}h(a)\Omega(a,t)-\mu P(a,t)-\lambda\frac{V}{H}h(a)\Omega(a,t-\tau)\xi$$

$$\frac{\partial W(a,t)}{\partial t}+\frac{\partial W(a,t)}{\partial a}=\lambda\frac{V}{H}h(a)\Omega(a,t-\tau)\xi-\mu W(a,t)$$

$$\frac{\partial M(a,t)}{\partial t}+\frac{\partial M(a,t)}{\partial a}=as\phi\left[W(a,t),k\right]W(a,t)-\gamma M(a,t)$$

$$\frac{\partial I(a,t)}{\partial t}+\frac{\partial I(a,t)}{\partial a}=W_T(a,t)-\delta I(a,t)$$

$$\frac{dL}{dt}=\lambda kg\int\pi(a)\left(1-f\left(M(a,t)\right)\right)da-\left(\sigma+\lambda\psi_1\right)L$$

$$L^{\square}=\frac{\lambda kg\int\pi(a)\left(1-f\left(M(a,t)\right)\right)da}{\sigma+\lambda\psi_1}$$

The above equations involve partial derivatives of four state variables (*P* - pre-patent worm load; *W* - adult worm load; *M* - microfilaria intensity; *I* - immunity to acquiring new infection due to the pre-existing total worm load where $W_T = W(a,t) + P(a,t)$). Given the faster time scale of infection dynamics in the vector compared to the human host, the infective L3-stage larval density in mosquito population is modelled by an ordinary differential equation essentially reflecting the significantly faster time-scale of the infection dynamics in the vector hosts. This allows us to make the simplifying assumption that the density of infective stage larvae in the vector population reaches a dynamic equilibrium (denoted by $L^*$) rapidly[1, 2, 5, 8, 9]. This basic coupled immigration-death structure of the model as well as its recent extensions has been extensively discussed previously[1-3, 5, 8, 9]. The effects of worm patency are captured by considering that at any time *t*, human individuals of age less than or equal to the pre-patency period, $\tau$, will have no adult worms or mf, and the rate at which pre-patent worms survive to become adult worms in these individuals at $a > \tau$ is given by $\zeta = \exp(-\mu\tau)$. The term enables us to account for the different establishment and development rates of the incoming L3-stage larvae as adult worms depending on the genus of mosquito vectors as expressed below:

$$f[\Lambda(a,t)] = \left[ \frac{2}{\left[1 + \frac{\Lambda(a,t)}{k}\left(1 - \exp\left[-\frac{r}{\kappa}\right]\right)\right]^{-k}} - \frac{1}{\left[1 + \frac{\Lambda(a,t)}{k}\left(1 - \exp\left[-\frac{2r}{\kappa}\right]\right)\right]^{-k}} \right]$$

for mosquitoes of *Anopheline* genus,

$$f[\Lambda(a,t)] = \left(1 + \frac{\Lambda(a,t)}{k}\left(1 - \exp\left[-\frac{r}{\kappa}\right]\right)\right)^{-k}$$

for mosquitoes of the Culicine genus.

In the above, $k\left[k_0 + k_{Lin} M(a,t)\right]$ is the shape parameter of the negative binomial distribution on the mf uptake whereas r and are respectively the rate of initial increase and the maximum level of L3 larvae. See Table S2 for the description of all the model parameters and functions.

**Table S2** – Description of EPIFIL model parameters and functions.

| Parameter | Definition (*units*) | Range | Refs |
|---|---|---|---|
| ***λ*** | Number of bites per mosquito (*per month*) | [5, 15] | 1, 2, 5, 10, 11 |
| ***τ*** | Pre-patency period | [1, 9] | 12 |
| ***s*** | Proportion of female worms | 0.5 | - |
| ***μ*** | The worm mortality rate (*per month*) | [0.008, 0.018] | 1, 2, 5, 13-16 |
| ***α*** | Production rate of microfilariae per worm (*per month*) | [0.25, 1.5] | 1, 2, 5, 17 |
| ***γ*** | The death rate of the microfilariae (*per month*) | [0.08, 0.12] | 1, 5, 15, 17 |
| ***g*** | Proportion of mosquitoes which pick up infection when biting an infected host | [0.251, 0.485] | 1, 5, 18 |
| ***κ*** | Maximum level of L3 given mf density | [3, 5] | 1, 5 |
| ***$k_0$*** | The basic location parameter of negative binomial distribution used in aggregation parameter ($k = k_0 + k_{Lin} M$) | [0.000036, 0.000775] | 1, 5, 19, 20 |
| ***δ*** | Immunity waning rate (*per month*) | [0.001, 0.01] | 1, 5 |
| ***V*** | Vector population size | [25000, 100000] | data |
| ***H*** | Human population size | data | data |
| ***$k_{Lin}$*** | The linear rate of increase in the aggregation parameter defined above | [0.00000024, 0.282] | 1, 5, 19, 20 |
| ***σ*** | Death rate of mosquitoes (*per month*) | [1.5, 8.5] | 1, 5, 20 |
| ***$\psi_1$*** | Proportion of L3 leaving mosquito per bite | [0.1, 0.8] | 17 |
| ***$\psi_2$*** | The establishment rate[1] | [0.00003, 0.00364] | 1, 2, 5, 21 |
| ***$H_{Lin}$*** | A threshold value used in *h(a)* to adjust the rate at which individuals of age *a* are bitten: linear rise from 0 at age zero to 1 at age $H_{Lin}$ in years. $h(a) = a/H_{Lin}$, for $a < H_{Lin}$; $h(a) = 1$ for $a \ge H_{Lin}$ | [240, 360] months | 1, 5, 9 |
| ***r*** | Gradient of mf uptake[2] | [0.04, 0.25] | 1, 5 |
| ***c*** | Strength of acquired immunity | [0.015, 0.025] | 1, 5 |
| ***$I_C$*** | Strength of immunosuppression[3] | [0.5, 5.5] | 1, 5 |
| ***$S_C$*** | Slope of immunosuppression function[4] (*per worm/month*) | [0.01, 0.20] | 1, 5 |
| | ***Intervention-related parameters*** | | |
| ***ω*** | Worm killing efficacy of drug (instantaneous) | dependent on drug regimen | 3 |
| ***ε*** | Microfilariae killing efficacy of drug (instantaneous) | dependent on drug regimen | 3 |
| ***$\delta_{reduc}$*** | Reduction in the worm's fecundity over a period of time *p* due to drug | dependent on drug regimen | 3 |
| ***p*** | A time period during which the drug remains efficacious in reducing the fecundity of the surviving adult worms | dependent on drug regimen | 3 |
| ***C*** | Percentage of the population administered the drug | data | data |
| ***$MBR_{VC}$*** | Vector control (VC) modifies $V/H$ ($= MBR/\lambda$) where $MBR_{VC} = MBR_0 \exp[qt]$, with $q<0$ for $\forall t$ when VC is implemented, otherwise $q=0$. | data and estimates | 19, 20 |

| Description | Mathematical expressions of the functions | Parameters | |
|---|---|---|---|
| Probability that an individual is of age *a* ***π(a)*** | $\pi(a) = A_0 \exp[-B_0 a]$ | Human age *a* in month, $A_0$ and $B_0$ estimated from country demographic data | 1, 5, 9 |
| Larvae establishment rate (modified by acquired immunity) ***Ω(a,t)*** | [illegible] | $\psi_1$ - proportion of L3 leaving mosquito per bite; $\psi_2$ - the establishment rate[1] | - |
| Adult worm mating probability ***φ(W,k)*** | $1-\left(1+\frac{W}{2k}\right)^{-(1+k)}$ | *k* – negative binomial aggregation parameter | 2, 5, 22 |
| Immunity to larval establishment ***$g_1$(I)*** | $\frac{1}{1+cI}$ | *c* – strength of immunity to larval establishment | 1, 5 |
| Host immunosuppression ***$g_2$($W_T$)*** | $\frac{1+I_C S_C W_T}{1+S_C W_T}$ | $I_C$ – strength of immunosuppression; $S_C$ – slope of immunosuppression | 1, 5 |

[1]The proportion of L3-stage larvae infecting human hosts that survive to develop into adult worms[2].
[2]The gradient of mf uptake *r* is a measure of the initial increase in the infective L3 larvae uptake by vector as *M* increases from 0[2, 9].
[3] The facilitated establishment rate of adult worms due to parasite-induced immunosuppression in a heavily infected human host
[4] The initial rate of increase by which the strength of immunosuppression is achieved as *W* increases from 0[23].
### Note MBR (monthly biting rate) serves as an input to initialize the model, measured as mosquito bites per person per month, the value of which may be obtained from entomological surveys conducted in study sites. In the absence of the observed MBR value, the model has been adapted to estimate it from the community-level mf prevalence data.

#### Model implementation

##### *Parameter selection and simulation procedure*

We employed the Bayesian Melding (BM) procedure to calibrate and estimate the LF models from field data, as outlined in detail in our previous work [2, 5-7]. Typically, we begin the procedure by first using the known or assignment of a uniform range for parameter values to generate distributions of parameter priors. We then randomly sample with replacement from these prior distributions to generate 200,000 parameter vectors, which are run using the annual biting rate (ABR) values, if given, for a site to generate model outputs. The model outputs are then melded with age-stratified mf prevalence data by calculating binomial log-likelihoods for each parameter vector. In the resampling step of the BM method, a Sampling-Importance-Resampling (SIR) algorithm is used to perform 500 draws with replacement from the pool of parameter vectors generated as above, with probabilities proportional to their relative log likelihood values. This step selects the parameter vectors which best describe the given mf age-prevalence data. These resampled parameter vectors are then used to generate distributions of variables of interest from the fitted model (eg. age-prevalence curves, worm breakpoints, and infection trajectories following treatments).

Here, we modified our standard Monte-Carlo BM framework for model discovery [2, 5-7, 24] to provide simulations for the chosen scenarios for two settings (one relevant for most of Africa, with Anopheles-driven transmission and annual treatment with ivermectin and albendazole (IA); and a second representing India-like populations, with Culex-driven transmission and treatment with diethylcarbamazine and albendazole (DA). The explicit aim was to generate at least N = 100,000 parameter vectors which resulted of overall mf prevalence values 1-60% at baseline in 2018. We first randomly sampled n = 100,000 parameter vectors from the assigned uniform parameter priors. We then simulated the endemic equilibrium given each parameter vector and calculated the predicted mf prevalence at baseline. Those parameter vectors whose outputs produced mf prevalence values between 1-60% were accepted while all others are rejected. This sampling and acceptance/rejection procedure was repeated until the total number of accepted parameter vectors (N) was greater than or equal to 100,000. The N posterior parameter vectors were then used to simulate the impacts of MDA and vector control interventions.

**Modeling intervention by mass drug administration**

Intervention by mass drug administration was modeled based on the assumptions that anti-filarial treatment with a combination drug regimen acts by killing certain fractions of the populations of adult worms and microfilariae instantly after the drug administration[25]. These effects are incorporated into the basic model by calculating the population sizes of worms and microfilariae as follows:

$$\left.\begin{aligned} W_f(a,t+dt) &= (1-\omega C)\,W_f(a,t) \\ W_m(a,t+dt) &= (1-\omega C)\,W_m(a,t) \\ M(a,t+dt) &= (1-\varepsilon C)\,M(a,t) \end{aligned}\right\} \quad \text{at time } t = T_{MDA_i}$$

where *dt* is a short time period since the *i*th MDA was administered. During this short time interval, a given proportion of adult worms and microfilariae are instantly removed. The parameters $\omega$ and $\varepsilon$ are drug killing efficacy rates for the two life stages of the parasite while the parameter *C* represents the MDA coverage. Apart from instantaneous killing of microfilariae, the drug continues to kill the newly reproduced mf by any surviving adult worms at a rate $\delta_{reduc}$ for a period of time, *p*. We model this effect as follows:

$$\frac{\partial M(a,t)}{\partial t} + \frac{\partial M(a,t)}{\partial a} = (1-\delta_{reduc} C)\,\alpha\,\phi(W_f(a,t), W_m(a,t))\,W_f(a,t) - \gamma M(a,t), \quad \text{for } T_{MDA_i} < t \le T_{MDA_i} + p$$

We simulated LF intervention by running the model with fixed values of $\omega$, $\varepsilon$, $\delta_{reduc}$, and *p* for MDA coverage levels given by the scenarios. The first MDA round was implemented in the model by affecting the population sizes of worms and microfilariae from the baseline estimates, and then the intervention is simulated forward in time for a number of years, with subsequent MDA rounds implemented annually or biannually.

**Modeling intervention by Vector Control**

In addition to MDA, we also modeled the added effect of long lasting insecticidal nets (LLINs) and indoor residual spray (IRS) as described previously [6]. The impact of LLINs with three main actions against mosquito biting was modelled: 1) deterrence from entering the home (efficacy $\eta_1$), 2) inhibition of their ability to feed on humans (efficacy $\eta_2$), and 3) killing them (efficacy $\eta_3$) [26,27] and the impact of IRS with two main actions against mosquito biting was modelled: 1) inhibition of their ability to feed on humans (efficacy $\eta_4$), and 2) killing them (efficacy $\eta_5$)[26,27]. To capture these effects, which decay over time as the larvicide efficacy declines exponentially at rate $\Lambda$, we adjust the term $V/H$ to be appropriately modified according to the population coverage for LLINs ($C_{LLIN}$) and IRS ($C_{IRS}$):

$$\frac{V}{H}\left(1-\eta_1 \exp\left(-\Lambda t\right) C_{LLIN}\right)\left(1-\eta_2 \exp\left(-\Lambda t\right) C_{LLIN}\right)\left(1-\eta_3 \exp\left(-\Lambda t\right) C_{LLIN}\right)$$
$$\left(1-\eta_4 \exp\left(-\Lambda t\right) C_{IRS}\right)\left(1-\eta_5 \exp\left(-\Lambda t\right) C_{IRS}\right)$$

#### LYMFASIM model description and methods

##### Description of the mathematical model

LYMFASIM[1,2] is a stochastic individual-based model for lymphatic filariasis (LF). It is a specific model variant within WORMSIM, a generalized framework for modelling transmission and control of helminth infections in humans[3,4]. LYMFASIM simulates the life histories of individual people and individual worms in a community, and the effects of interventions (e.g. mass drug administration, integrated vector management, bednet use) on transmission and morbidity, while taking into account of the human demography and the complexities of helminth transmission. The model has been described elsewhere and has been applied to support decision making on control and elimination of lymphatic filariasis in different settings[1,2,5-12].

Mass drug administration (MDA) is simulated by specifying the year and month in which treatment takes place, the efficacy of the applied treatment regimen, the achieved coverage level, and compliance patterns. LYMFASIM assumes that a fraction of the population never participates in MDA (e.g. systematic refusal, related to chronic illness). In addition, LYMFASIM allows the relative compliance to vary between age and sex groups; this mechanism captures transient contra-indications for MDA (e.g. exclusion of young children and pregnant women) and other age- and sex-related behavioral factors driving participation in MDA. Lastly, each individual has a personal inclination to participate in MDA, which is considered as a lifelong property. A stochastic process eventually defines for each individual whether they are treated in a given round, depending on the calculated probability.

##### Parameter quantification and simulation methods for this study

We previously derived model quantifications for simulating LF transmission by *Culex quinquefasciatus* in India[2] and by Anopheles species in Africa[8]. These model quantifications differ in the age-structure of the human population and the density dependence in the L3 yield from a blood meal in mosquitoes. For India, we originally developed several model variants[2]: two model variants assumed that people would acquire some form of acquired immunity against infection during their lifetime, as a consequence of being exposed to various stages of infection; one model variant assumed no immunity. The variants with immunity fitted best to the age-specific data that showed a decline in prevalence in older age group. Of these, the 'anti-L3' variant (with immunity triggered by incoming L3 larvae and reducing the probability for these larvae to survive and develop successfully into adult worms) has been used most in later studies[5-7] as it seemed most consistent with external antigenaemia prevalence data. As a result of strong acquired immunity, the range of mf prevalences that can be simulated with this model is relatively narrow.

A review of age-prevalence patterns[13] has casted doubt about the assumed role of acquired immunity. In view of these doubts, acquired immunity was not considered to play a role in the Africa model[8]. We now also revert to the model variant without acquired immunity for India. As shown by Subramanian, exclusion of immunity requires some changes in other parameters, notably the success ratio (i.e. the proportion of incoming L3 larvae that survives and develops successfully into an adult worm) and the assumed variation in exposure between individuals[2]. In the anti-L3 immunity model variant, the success ratio is down regulated as a consequence of acquired immunity. In a model without such immunity, we therefore need to assume a lower value. Also, to simulate the relatively low pre-control mf prevalence levels that are common in India without acquired immunity, we need to assume more heterogeneity in exposure with many individuals having a low exposure and a small group of people being highly exposed to mosquito bites. Subramanian proposed a model in which part of the simulated population does not participate in transmission. We take a slightly different approach in this paper, combining relatively low average monthly biting rates (mbr) with high exposure heterogeneity. The monthly biting rate and parameter describing the extent of exposure heterogeneity (i.e. the shape parameter of the Gamma distribution that describes exposure heterogeneity) are varied according to a predefined parameter space as defined below.

Parameter values are listed in Table S3. Assumptions and parameters related to control strategies and treatment efficacy are listed in Table S4. The monthly biting rate (mbr), exposure heterogeneity parameter (k) and external force-of-infection (foi) were varied according to the density plots in Figure S5, in order to generate simulations across a wide range of mf prevalences at baseline, measured in the total population (all ages).

**Version and code availability**

For this paper, we used WORMSIM version 2.58Ap27, available from
https://gitlab.com/erasmusmc-public-health/wormsim.previous.versions/-/blob/master/wormsim-2.58Ap27.zip.

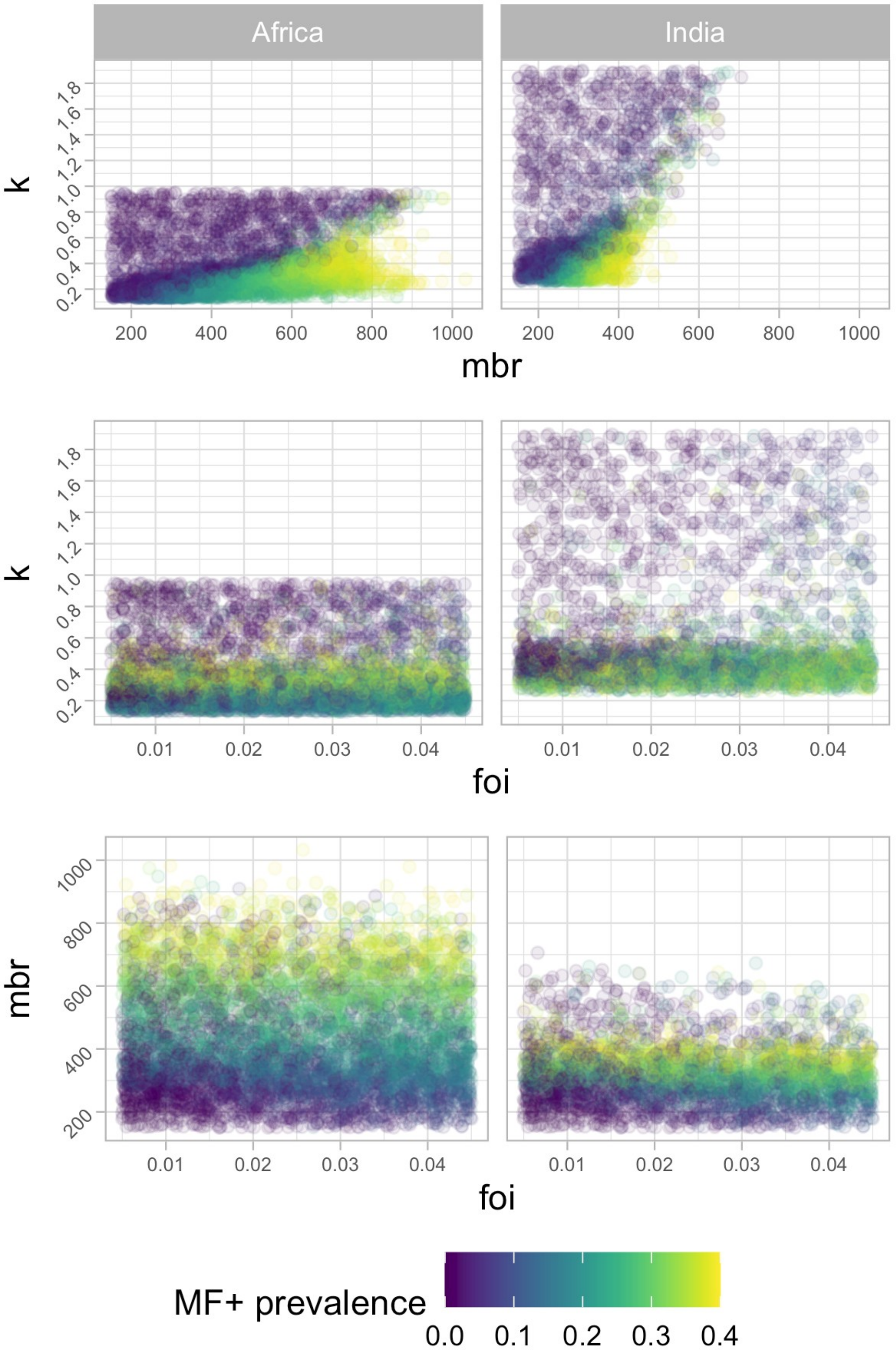

**Figure S5.** Density plots illustrating the parameter space areas for the monthly biting rate (mbr), parameter describing exposure heterogeneity in the population (k) and external force of infection (foi), as used to generate simulations with baseline mf prevalence in the range from 0 to 40%, for the Africa and India model variants.

Table S3 – LYMFASIM input: probability distributions, functions and parameter values for simulating transmission of bancroftian filariasis by Anopheles mosquitoes in Africa or Culex in India

| **Parameter description** | **Anopheles model** | | **Culex model** | | **Source / remarks** |
|---|---|---|---|---|---|
| **Human demography** | | | | | |
| Cumulative survival, by age | Age | Survival | Age | Survival | Anopheles: Modified from ref[8]¸to match the population composition in Ethiopia; Culex: Fixed as in[2] |
| (upper limit age category) | 0 | 1 | 0 | 1 | |
| | 5 | 0.800 | 5 | 0.904 | |
| | 15 | 0.790 | 10 | 0.895 | |
| | 20 | 0.755 | 15 | 0.888 | |
| | 25 | 0.737 | 20 | 0.879 | |
| | 30 | 0.723 | 25 | 0.864 | |
| | 35 | 0.654 | 30 | 0.849 | |
| | 40 | 0.605 | 40 | 0.812 | |
| | 45 | 0.560 | 50 | 0.756 | |
| | 50 | 0.506 | 90 | 0 | |
| | 60 | 0.487 | | | |
| | 70 | 0.305 | | | |
| | 80 | 0.155 | | | |
| | 99 | 0.000 | | | |

| Parameter description | Anopheles model | | Culex model | | Source / remarks |
|---|---|---|---|---|---|
| Fertility rate per woman, by age | Age | Fertility rate | Age | Fertility rate | Anopheles: fixed, as in [8]; Culex: fixed as in [2] |
| (upper limit age category) | 0 | 0 | 0 | 0 | |
| | 5 | 0 | 5 | 0 | |
| | 15 | 0 | 10 | 0 | |
| | 20 | 0.116 | 15 | 0 | |
| | 25 | 0.230 | 20 | 0.075 | |
| | 30 | 0.245 | 25 | 0.254 | |
| | 35 | 0.207 | 30 | 0.222 | |
| | 40 | 0.147 | 40 | 0.096 | |
| | 45 | 0.077 | 50 | 0.013 | |
| | 50 | 0.031 | 90 | 0 | |
| | 60 | 0 | | | |
| | 70 | 0 | | | |
| | 80 | 0 | | | |
| | 99 | 0 | | | |
| Initial population | age | Male/females | Age | Males/females | N/A |
| | 5 | 42/42 | 5 | 20/20 | |

| Parameter description | Anopheles model | | Culex model | | Source / remarks |
|---|---|---|---|---|---|
| | 15 | 63/63 | 10 | 17/17 | |
| | 20 | 26/26 | 15 | 15/15 | |
| | 25 | 22/22 | 20 | 15/15 | |
| | 30 | 20/20 | 25 | 22/22 | |
| | 35 | 17/17 | 30 | 20/20 | |
| | 40 | 14/14 | 40 | 15/15 | |
| | 45 | 11/11 | 50 | 13/13 | |
| | 50 | 9/9 | 90 | 13/13 | |
| | 60 | 14/14 | | | |
| | 70 | 9/9 | | | |
| | 80 | 3/3 | | | |
| | 99 | 1/1 | | | |
| Maximum population size | Varied according to population size distribution defined elsewhere in this manuscript | | Varied according to population size distribution defined elsewhere in this manuscript | | Figure S1 |
| Proportion removed when maximum population size is reached | 5% | | 5% | | N/A |
| **Exposure** | | | | | |

| Parameter description | Anopheles model | Culex model | Source / remarks |
|---|---|---|---|
| External force-of-infection at start of burn-in period | 2 | 2 | N/A |
| Duration of external force-of-infection at start of burn-in period | 2 years | 2 years | N/A |
| External force-of-infection (foi) during the simulation | Varied between runs, as specified in figure S1; assumed to remain constant during the entire simulated period | Varied between runs, as specified in figure S1; assumed to remain constant during the entire simulated period | N/A |
| Average mosquito biting rate for adult men (mbr), for a relative biting rate of 1 | Varied between runs, as specified in figure S1 | Varied between runs, as specified in figure S1 | N/A |
| Seasonal variation in biting rate | No | No | N/A |
| Variation in exposure by age (no difference assumed between sexes) | 0 at birth, linearly increasing to reach 1 at the age of 20 and constant at 1 from this age onwards | 0.26 at birth, linearly increasing to reach 1 at the age of 19.1 and constant at 1 from this age onwards | Anopheles: slightly adjusted from [2] for Africa[8]; Culex: estimated by fitting to data[2]; |
| Probability distribution describing variation in the individual exposure index, due to personal factors (fixed through life) given age and sex | Varied between runs, as specified in figure S1 | Varied between runs, as specified in figure S1 | Gamma distribution is assumed as in[2,8]; shape/rate parameter varied |

| Parameter description | Anopheles model | Culex model | Source / remarks |
| --- | --- | --- | --- |
| **Parasite dynamics within host** | | | |
| Success ratio | 0.00088 | 0.00058 | Previously estimated by fitting to data[8,2] (no immunity model in [2]) |
| Anti-L3 immunity | Not included in the model, by assuming that the strength and duration of the immunological memory are zero | Not included in the model, by assuming that the strength and duration of the immunological memory are zero | Assumed[8] |
| Anti-fecundity immunity: | Not included in the model, by assuming that the strength and duration of the immunological memory are zero | Not included in the model, by assuming that the strength and duration of the immunological memory are zero | Assumed[8] |
| Average worm lifespan | 10 years on average; varied according to a Weibull distribution with shape 2 | 10.2 on average; varied according to a Weibull distribution with shape 2 | Previously estimated by fitting to data[2] |
| Duration of immature stage of the parasite in human host | Constant, 8 months | Constant, 8 months | Fixed, based on [14] |
| No. of Mf produced per female parasite per month per 20 ml peripheral blood in the absence of immune reactions and in the presence of at least 1 male worm | 0.58 | 0.606 | Previously estimated by fitting to data[2] |
| Monthly survival of the microfilariae, fraction | 0.9 | 0.9 | Fixed, based on[15] |
| Association between worm age and | mf production independent of worm | mf production independent of worm | Assumed |

| Parameter description | Anopheles model | | Culex model | | Source / remarks |
|---|---|---|---|---|---|
| mf production rate | age | | age | | |
| Polygamy (all female worms produce mf in the presence of at least one male worm) | Yes (male potential 1000) | | Yes (male potential 1000) | | Assumed |
| Mating cycle (number of months a female can produce mf with one insemination) | 1 | | 1 | | Assumed |
| **Uptake of infection by the vector** | | | | | |
| Functional relationship, where L3 = the average number of L3 larvae developing in mosquitoes and M = the mf density in human blood as counted in a 20µL bloodsmear | $L3 = a$ | | $L3 = c + \frac{aM}{1 + aM/(b-c)}$ | | Fixed, based on [8,16] |
| | a | 1.666 | a | 0.089 | |
| | b | 0.027 | b | 6.6 | |
| | c | 1.514 | c | 0 | |
| Transmission probability (v), fraction of the L3 larvae, resulting from a single blood meal, that is released by a mosquito | 0.1 | | 0.1 | | Fixed, as in[8] |
| **Other** | | | | | |

| Parameter description | Anopheles model | Culex model | Source / remarks |
|---|---|---|---|
| Start year of simulation period (burn-in period runs from this start-year to the moment of first intervention) | 1850 | 1850 | N/A |
| **Surveillance** | | | |
| Timing of surveys | Yearly, at the start of a calendar year from 2018-2042 onwards (preceding mass treatment, when it occurs in the same year) | Yearly, at the start of a calendar year from 2005-2030 onwards (preceding mass treatment, when it occurs in the same year) | N/A |
| Volume of blood examined for mf | 60 µL | 60 µL | N/A |
| Variability in observed number of mf in one 20 µl blood smear | Negative binomial distribution with k=0.33 | Negative binomial distribution with k=0.33 | Previously estimated for 20 µL blood by fitting to data[2] |
| Variation between worms in their contribution to measured mf count (dispersal factor) | Constant (no variation) | Constant (no variation) | Assumed |
| **Morbidity** | | | |
| not applicable | | | |
| no excess mortality due to disease | | | |

**Table S4** – LYMFASIM assumptions related to interventions scenarios: MDA and bednet use

| Parameter | Values | | Source |
|---|---|---|---|
| **Bednet use** | Not included | | |
| **Mass treatment** | | | |
| Timing of mass treatment rounds, and coverage and fraction excluded per round | *See main text for an overview of scenarios considered* | | N/A |
| *Relative compliance by age and sex for treatment with ivermectin + albendazole* | | | |
| age-group | Males | Females | Based on unpublished data from the Onchocerciasis Control Programme (OCP) data on compliance with ivermectin mass treatment for onchocerciasis |
| 0-4[b] | 0[a] | 0[b] | |
| 5-9 | 0.75 | 0.75 | |
| 10-14 | 0.80 | 0.70 | |
| 15-19 | 0.80 | 0.74 | |
| 20-29 | 0.70 | 0.65 | |
| 30-49 | 0.75 | 0.70 | |
| 50+ | 0.80 | 0.75 | |
| *Relative compliance by age and sex for treatment with diethylcarbamazine + albendazole, and for ivermectin + diethylcarabamazine + albendazole* | | | |
| age-group | Males | Females | Adapted from OCP data on ivermectin treatment (see above) |
| 0-1[b] | 0[c] | 0[c] | |
| 2-9 | 0.75 | 0.75 | |
| 10-14 | 0.80 | 0.70 | |
| 15-19 | 0.80 | 0.74 | |
| 20-29 | 0.70 | 0.65 | |
| 30-49 | 0.75 | 0.70 | |
| 50+ | 0.80 | 0.75 | |
| **Drug treatment** | | | |
| Fraction malabsorption (no effect) | 0% | | |
| Efficacy ivermectin + albendazole | | | |
| Proportion of adult worms killed per treatment, average | 35% | | |
| Duration of temporary reduction in female reproductive capacity, average | 9 months[c] | | |

[a] Children under 5 are assumed to be exempted from ivermectin + albendazole treatment

[b] Children under 2 are assumed to be exempted from diethylcarbamazine +albendazole treatment; children between 2-5 are treated, although they don't receive ivermectin in case of triple drug treatment.

[c] Assuming complete absence of mf production during this period and immediate resumption of

| Parameter | | Values | Source |
|---|---|---|---|
| | Permanent reduction in female worm reproductive capacity, average | 0% | |
| | Variability in effect of treatment on adult worms | Not applicable (assumed constant, 1) | |
| | Fraction of mf surviving per treatment | 1% | |
| Efficacy diethylcarbamazine + albendazole | | | |
| | Proportion of adult worms killed per treatment, average | 55% | |
| | Duration of temporary reduction in female reproductive capacity, average | 6 months | |
| | Permanent reduction in female worm reproductive capacity, average | 0% | |
| | Variability in effect of treatment on adult worms | Not applicable (assumed constant, 1) | |
| | Fraction of mf surviving per treatment | 5% | |
| Efficacy ivermectin + diethylcarbamazine + albendazole | | | |
| | Proportion of adult worms killed per treatment, average | 55% | |
| | Duration of temporary reduction in female reproductive capacity, average | N/A | |
| | Permanent reduction in female worm reproductive capacity, average | 100% | |
| | Variability in effect of treatment on adult worms | Not applicable (assumed constant, 1) | |
| | Fraction of mf surviving per treatment | 0% | |

mf production thereafter

#### TRANSFIL model description and methods

##### Description of the mathematical model

The mathematical model of lymphatic filariasis (LF) transmission TRANSFIL is a stochastic individual-based model of LF infection in human populations. A full model description is given in Irvine *et al.*[1] and more recently in Michael *et al.*[2], so here we provide a brief summary of the model development. TRANSFIL is a stochastic individual-based model, simulating worm burden, microfilaraemia and other demographic parameters relating to age and risk of exposure. Humans are modelled individually, with their own male and female worm burden. The concentration of mf in the peripheral blood is modelled for each individual and increases according to the number of fertile female worms as well as decreasing at a constant rate.

The total mf density in the population contributes towards the current density of L3 larvae in the human-biting mosquito population, where the distribution of L3 amongst the human-biting mosquito population is completely homogeneous. An empirically derived relationship is used for the uptake of mf by a mosquito, where both *Culex* and *Anopheles* uptake curves are implemented depending on setting (see Irvine *et al.*[1]). The model dynamics are therefore divided into the individual human dynamics, including age and turnover; worm dynamics inside the host; microfilariae dynamics inside the host and larvae dynamics inside the mosquito.

##### Model implementation

We performed simulations for LF endemic settings in India and Africa, accounting for different population sizes, possible range of precontrol mf prevalence levels (prevalence range 0 to 60% in the scenarios considered) and regional differences in local vector species and standard treatment regimens (*Culex* and DA in India, *Anopheles* and IA in Africa). More specifically, in order to generate the required range of mf prevalences in individuals above 5 years of age, we varied three parameters of the model, the vector to host ratio (V/H), the average population bite risk (k) and the importation rate (Imp), using parameter sets from a range of plausible values based on previously analysed data[1,2,3]. The graphical representation of the values is shown in Figures S6-S8.

For stochastic models it is essential that an importation rate is included, otherwise the equilibrium distribution (steady state) that is used as the starting point of the simulations can potentially converge to the degenerate distribution where no-one is infected. The importation rate does not need to be large, in fact it should not be driving the infection. For this LF study we used a random number drawn from a uniform distribution with minimum 0 and maximum 0.00025 (max 2.5/10000 infections per month). The interventions reduce the prevalence over time, and therefore as years pass, the importation rate decreases in proportion to the reduction in prevalence seen in some pilot simulations. More specifically, we produced 2000 simulations for each scenario with constant importation rate over the years (including the years of MDA). Then, for our main set of simulations, we adjust the importation rate according to how the prevalence changed in our pilot runs after the intervention was applied (for example see Figure S9).

Finally, compliance between rounds of MDA is modelled based on the paper by Griffin *et al.*[14], following the description in Dyson *et al.*[15] and previous implementation of the model[2]. A summary of all model parameters is available in Table S5.

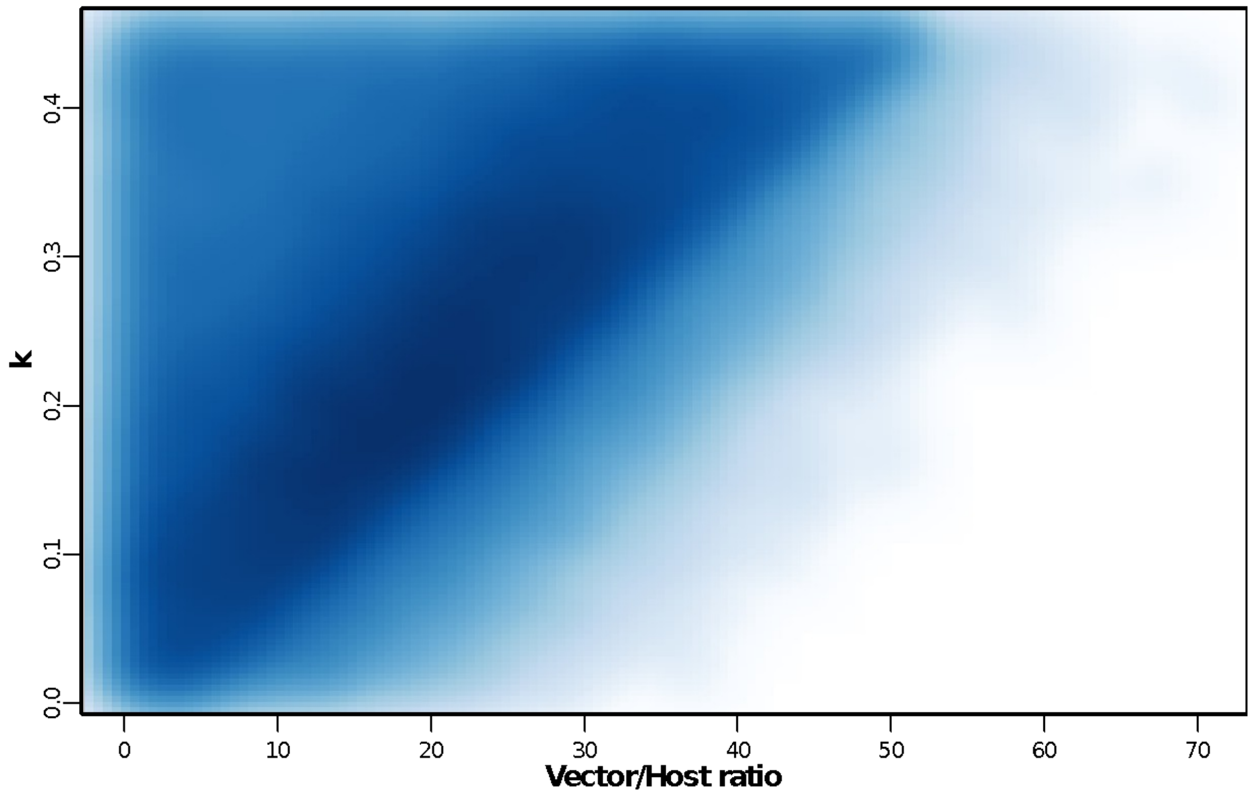

**Figure S6** – Vector to Host ratio against aggregation parameter k. The density plot indicates the parameter space areas from the simulations that were in the 0 to 60% mf prevalence range at baseline. Dark blue areas denote more common parameter values.

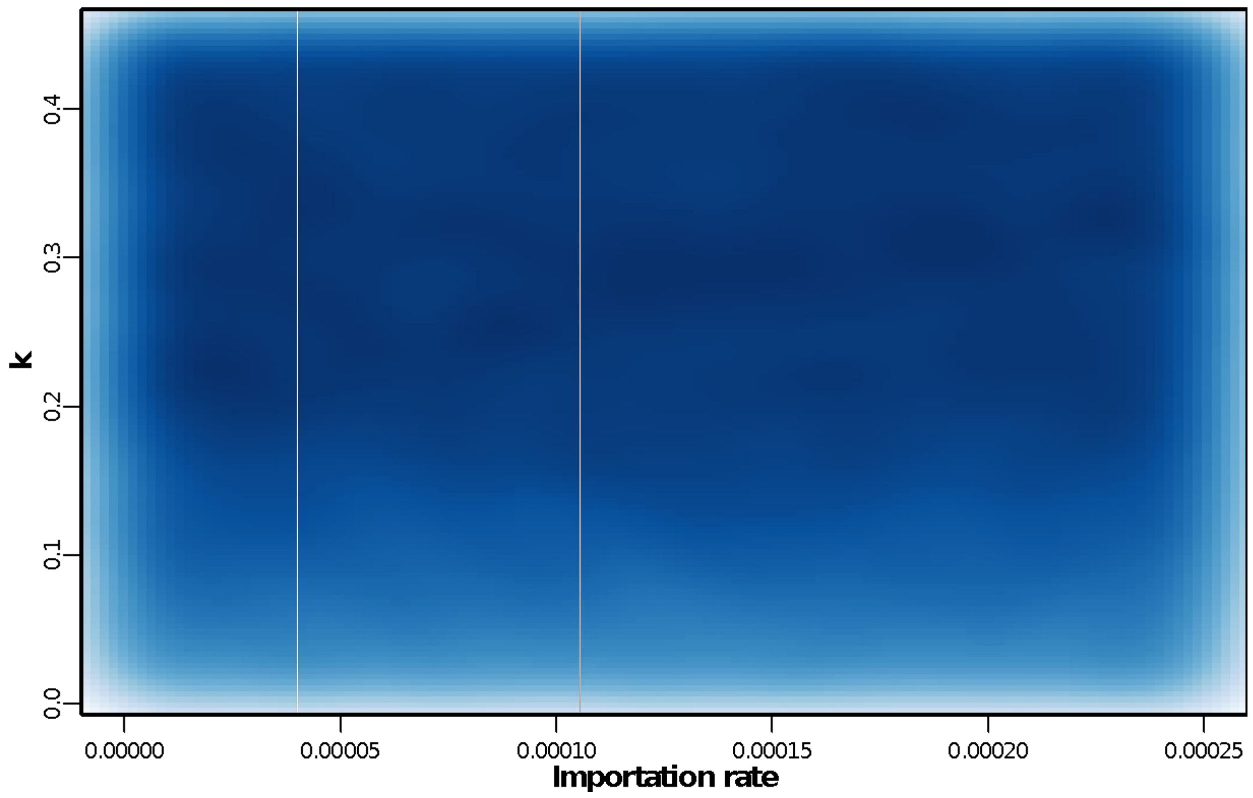

**Figure S7** – Importation rate against aggregation parameter k. The density plot indicates the parameter space areas from the simulations that were in the 0 to 60% mf prevalence range at baseline. Dark blue areas denote more common parameter values.

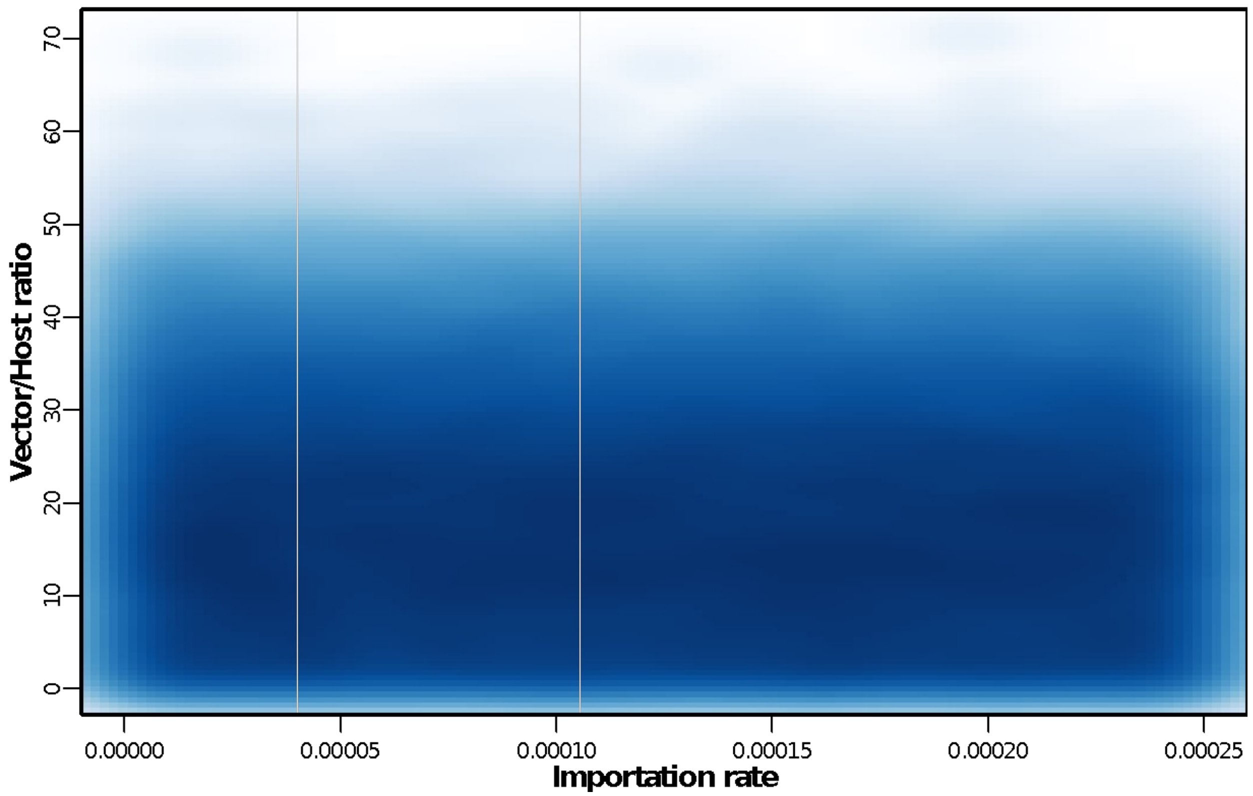

**Figure S8** – Importation rate against Vector to Host ratio. The density plot indicates the parameter space areas from the simulations that were in the 0 to 60% mf prevalence range at baseline. Dark blue areas denote more common parameter values.

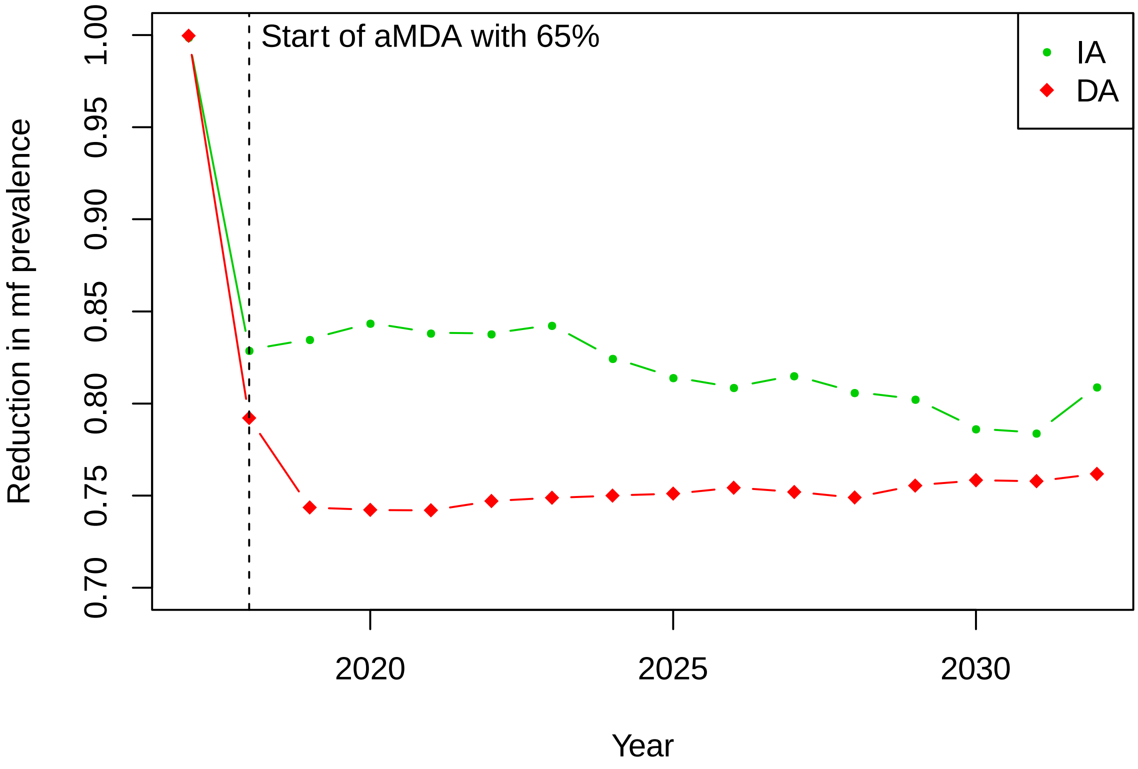

**Figure S9** – Reduction in mf prevalence over the years seen in the pilot simulations. The first intervention took place in 2018, annual MDA with 65% coverage (IA green and DA red), and was applied for 15 rounds with no interruptions.

**Table S5** – Description the basic LF model parameters.

| **Parameter symbol** | **Definition** | **Value** | **Source** |
|---|---|---|---|
| λ | Number of bites per mosquito | 10 per month | [4,5] |
| V/H | Ratio of number of vectors to hosts | Varied | Input |
| $\alpha_{max}$ | Age at which exposure to mosquitoes reaches its maximum level | 20.0 | [6] |
| $\psi_1$ | Proportion of L3 leaving mosquito per bite | 0.414 | [7] |
| $\psi_2$ | Proportion of L3 leaving mosquito that enter host | 0.32 | [8] |
| $s_2$ | Proportion of L3 entering host that develop into adult worms | 0.00275 | [9,10] |
| μ | Death rate of adult worms | 0.0104 per month | [11] |

| | | | |
|---|---|---|---|
| $\delta$ | Production rate of mf per worm | 0.2 per month | [7] |
| $\zeta$ | Death rate of mf | 0.1 per month | [7,12] |
| g | Proportion of mosquitoes which pick up infection when biting an infected host | 0.37 | [13] |
| $\sigma$ | Death rate of mosquitoes | 5 per month | [8] |
| k | Aggregation parameter of individual exposure to mosquitoes | Varied | Input |
| $h(\alpha)$ | Parameter to adjust rate at which individuals of age $\alpha$ are bitten | Linear from 0 to 10, with maximum of 1 | [9] |
| Imp | Importation rate | Varied | Input |
